# Evaluating Standards-Based Audit for Quality Improvement in Maternal and Newborn Care: Evidence from a multi-country implementation study

**DOI:** 10.1101/2025.11.20.25340669

**Authors:** Fiona M. Dickinson, Lydia Mwanzia, Hauwa Mohammed, Leonard Katalambula, Salma Abdi Mahmoud, Irene Nyaoke, Onesmus Maina, Martin Eyinda, Alice Norah Ladur, Helen Allott, Uzoh Egere, Charles Ameh

## Abstract

**Background:** Improving maternal and newborn health outcomes in low- and middle-income countries requires not only increased service coverage but also enhanced quality of care. Standards-based audits offer a structured, evidence-based approach to quality improvement; however, their implementation and effectiveness across diverse low- and middle-income country contexts remain underexplored.

**Objectives:** To assess the implementation and effectiveness of a quality improvement training programme using standards-based audit methodology across health facilities in Kenya, Nigeria, and Tanzania, and to evaluate its contribution to improvements in the quality of maternal and newborn care.

**Methods:** Between 2020 and 2024, 895 healthcare workers from 211 facilities, were trained using traditional and blended learning methods. One hundred and twenty-eight facilities were followed up, assessing the implementation of quality improvement. Facilities identified context-specific care gaps, audited baseline performance, implemented targeted interventions, and conducted follow-up audits to improve care.

**Results:** All facilities appointed a quality improvement champion, identified a care gap, and audited performance against an appropriate standard (n=137 audits/128 facilities). The average baseline audit scores across all three countries were 38.8%, with follow-up scores increasing to 77.4%, indicating a mean improvement of 38.2%. Over half (54%) of the facilities met or exceeded their improvement targets.

Key challenges include unrealistic target setting, resource constraints, and context-specific issues (staff shortages and supply disruptions). The approach was feasible across all facility levels. Contextualised standard selection enhanced local relevance and ownership of interventions, while the blended learning model increased the accessibility and cost-efficiency of the training.

**Conclusion:** Standards-based audit and quality improvement training is a scalable and effective approach to improve maternal and newborn care in low- and middle-income countries. The findings of this multi-country study support the broader adoption of low-tech, standards-driven methods and highlight the need for further research into cost-effectiveness and long-term health outcomes to inform national scale-up strategies.

## Introduction

While the coverage of maternal and newborn health services is important, to improve health outcomes, the quality of care provided also needs to be of a high standard. Tuncalp et al (2015) describe poor quality care as “a paramount roadblock” to efforts to prevent maternal and neonatal mortality and morbidity. Globally, over 700 women (WHO, 2025a) and approximately 6,500 newborns (WHO, 2024) lose their lives daily, during and following pregnancy and childbirth. In 2023, sub-Saharan Africa accounted for approximately 70% of all maternal deaths and 57% of under-5 deaths globally and 57% of under-5 deaths. Most maternal deaths are due to conditions such as uncontrolled bleeding, high blood pressure, infection, and abortion (WHO, 2025a), which are preventable with good quality care.

Good quality care has been characterised in various ways but should include the use of appropriate evidence-based, clinical, and non-clinical interventions (Tuncalp et al., 2015). One strategy for addressing the need to improve care quality is through the use of standards-based audits (StBA). A clinical audit has been defined as “part of the continuous quality improvement process”, consisting of “measuring a clinical outcome or process against well-defined standards, established using the principles of evidence-based medicine” (Esposito & Canton, 2014). A study exploring the effectiveness of StBA related to emergency obstetric care in Malawi found significant improvements in the quality of care provided following the implementation of the intervention (White et al., 2024).

The Emergency Obstetric and Quality of Care Unit at the Liverpool School of Tropical Medicine (LSTM) developed a suite of training courses for healthcare providers, including emergency obstetric care, antenatal and postnatal care, and quality improvement (QI). The initial QI course comprised a 5-day, face-to-face (F2F) training, facilitated by three master trainers, and supported with a printed training manual.

However, the Covid-19 pandemic acted as a catalyst for the repackaging of the training course to make it suitable for delivery using a flipped classroom, blended learning (BL) approach. Flipped classroom refers to a student-centred approach which inverts or ‘flips’ the traditional ‘teacher’ led approach. The flipped approach provides students with learning materials and tasks, which they are then required to work through independently, prior to more traditional face-to-face sessions, to assess and address any gaps in their learning. This entails that students become active participants in their own development. (Ettien & Toure, 2023)

The BL approach incorporated in the QI course combined self-directed online learning (SDL), group online learning, and group F2F learning. The SDL used the World Continuing Education Alliance online platform and provided 10 sets of pre-recorded lectures and learning activities covering the theoretical aspects of QI. Additionally, three online, real-time, group learning activities, were held, lasting approximately 2.5 hours each and supported by experienced facilitators. They covered a combination of theoretical and practical activities to ensure that participants understood the QI approach and its implementation. The final aspect of the course involved a 1-day, face-to-face (F2F) group learning activity, reinforcing the practical aspects of QI and supporting participants in developing a QI plan for their own workplaces. Additional support materials included participant workbooks containing templates and worked examples, a detailed facilitator’s e-manual, and orientation sessions for new facilitators. The BL approach had the added benefit of making the course accessible to more participants and being cost-efficient in a manner that has been found to work well for other courses (Ladur et al., 2025; Vallee et al., 2020).

The LSTM QI course, both in its traditional F2F and revised BL formats, provided healthcare workers with the necessary tools to evaluate and improve the quality of care provided in facilities using StBA (Figure 1).

**Figure 1.**
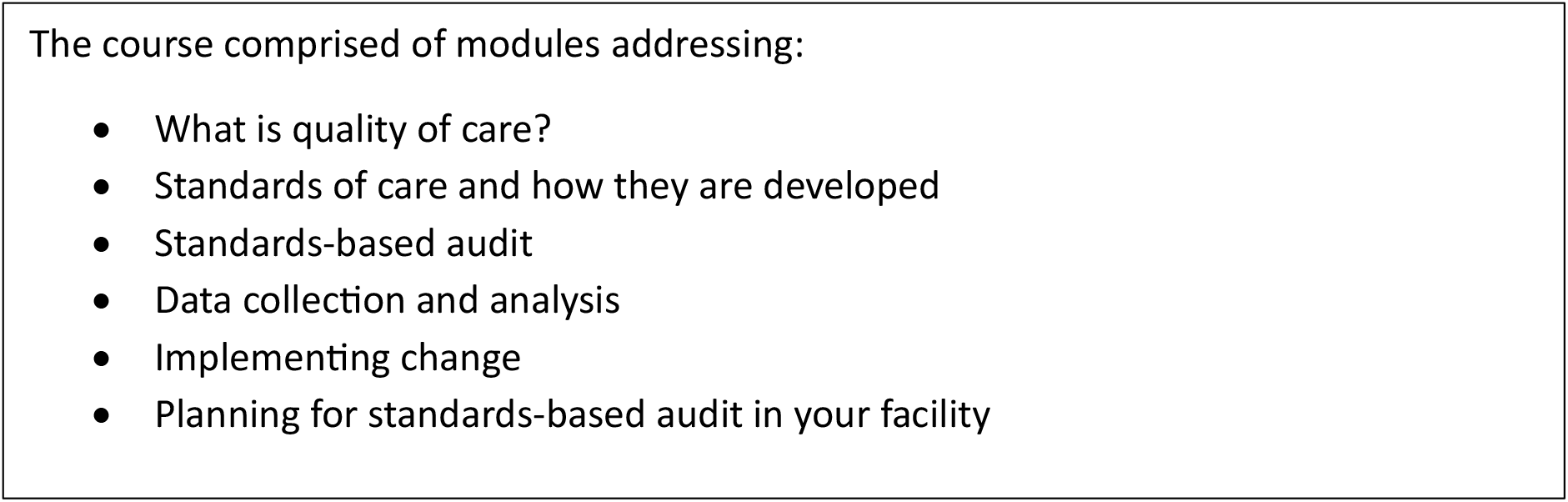
Summary of QI course content

The method of conducting standards-based audits taught in the LSTM training (Figure 2) involved the identification of a clinical problem resulting in poor outcomes within a health facility and an appropriate standard relating to the specific problem. Standards were obtained from national or international guidelines or developed locally based on current research and best practices. A baseline assessment was conducted in the study facilities with a random sample of women against the standard (M1), and a target for quality improvement was set. The precise level of the target was dependent on the baseline level and the realistic likelihood of achieving it. Facilities then developed an intervention to address the problem and implemented it, conducting a follow-up assessment (M2) against the same standard after three months. If the target was achieved, facilities selected another problem and standard to address, while facilities that were unable to achieve their target explored the reasons for why it was not achieved and repeated the process for a second cycle.

**Figure 2.**
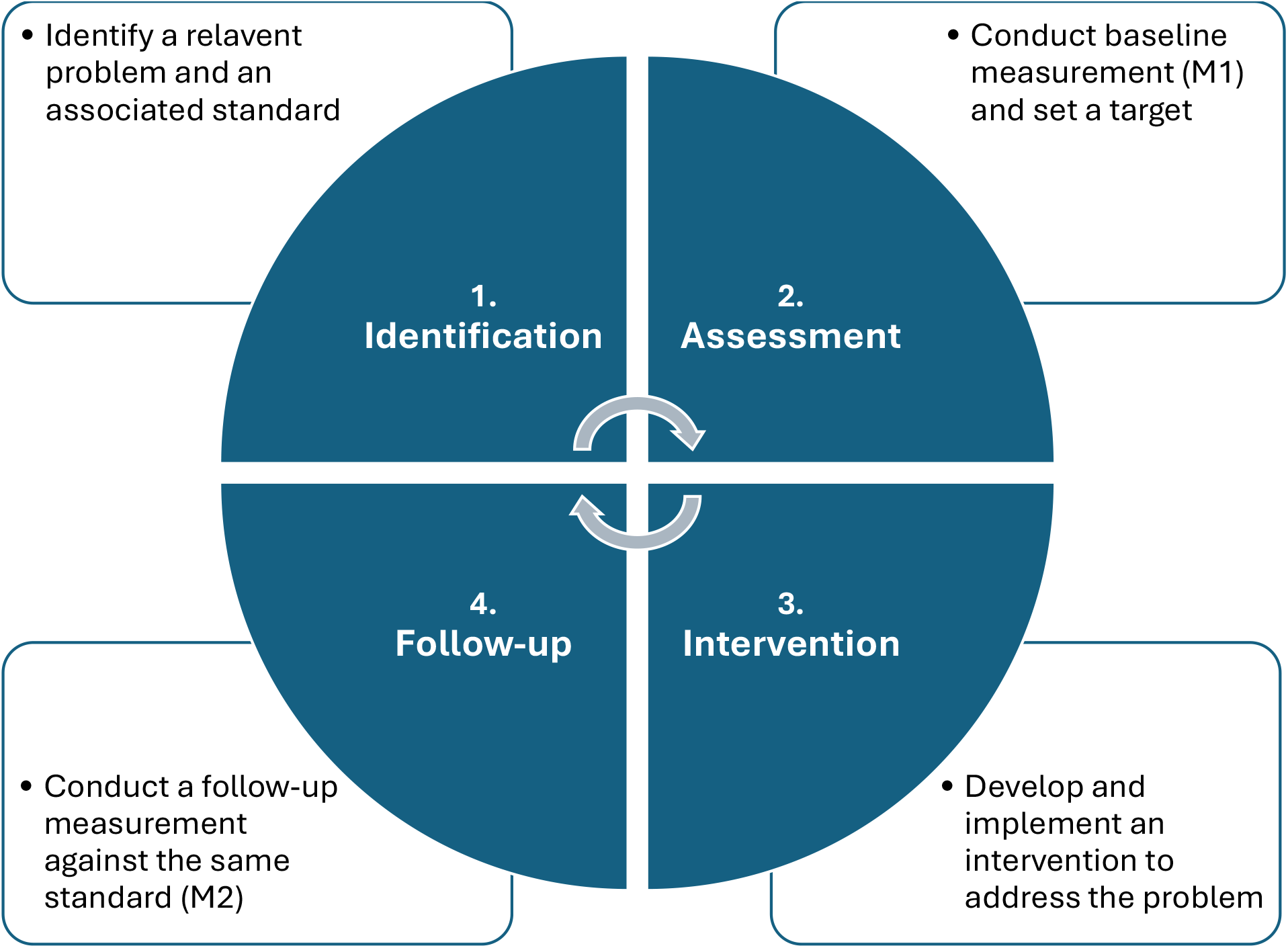
Standards-based audit methodolog

To our knowledge, there are no published large-scale evaluations of QI training programmes incorporating StBA methodology, implemented across diverse maternity care settings of differing levels, in multiple low- and middle-income countries (LMICs). This study aimed to assess the implementation of StBA following QI training and evaluate the effectiveness of a novel QI training program in improving the quality of care in selected facilities in Nigeria, Kenya, and Tanzania.

## Methods

This observational study sought to assess the effect of QI training on the implementation and outcomes of StBA in selected healthcare facilities and is reported using SQUIRE 2.0 guidelines. QI training was conducted in the three countries included in the Global Fund programme: Kenya, Nigeria, and Tanzania (mainland (ML) and Zanzibar) between July 2020 and April 2024. The World Bank (2025) classifies these countries as lower-middle income. In total, 895 healthcare providers from 211 government health facilities were trained in QI methods across 28 courses (Table 1).

**Table 1.** Summary of QI trainings in Kenya, Nigeria & Tanzania.

|  | Kenya | Nigeria | Tanzania ML | Tanzania Zanzibar | Total |
| --- | --- | --- | --- | --- | --- |
| Number of QI courses | 9 | 9 | 8 | 2 | 28 |
| Number of HCW trained in QI | 323 | 330 | 188 | 54 | 895 |
| Number of facilities with staff trained in QI | 61 | 120 | 20 | 10 | 211 |
| Number of facilities followed up | 61 | 37 | 20 | 10 | 128 |

In Nigeria, the Ministry of Health (MoH) selected the facilities, targeting high-volume facilities in Oyo and Kaduna states. Participants were selected by the state-level ministries of health and primary health care development agencies. LSTM had no influence on who was selected for training, and all trainees came from ANC/PNC clinics.

A similar process was followed in Kenya, where high-volume facilities were selected from three counties (Garissa, Vihiga, and Uasin Gishu) based on the number of births, maternal and perinatal deaths, and ANC and PNC visits per facility, with additional input from County Health Management Teams.

In Tanzania, facilities were selected using a stratified random sampling method based on facility level and geographical location from Dodoma, Unguja, and Pemba regions. All facilities were government-run and included dispensaries, health centres, and hospitals.

As part of the training, the QI champions developed a QI implementation plan for their respective facilities. Following training, the QI champions disseminated the plan and StBA methods to their respective facilities,prompting the formation or revitalisation of QI teams, which, alongside the QI champions, led the audit process. QI champions and teams were supported by the LSTM through the provision of basic ANC and PNC equipment and sharing QI materials and advice via country-specific instant messaging groups.

Periodic follow-up monitoring was conducted by members of the LSTM staff and partners based in each country as part of the Global Fund programme. These recorded the extent to which the StBA process had been implemented, scores obtained for M1 and M2, facilities actions following M2, and challenges faced during the process. Data were recorded using a standardised spreadsheet. The scores for M1 and M2 were derived from the number of women for whom the standard of care had been met, out of an overall sample audited (typically 20 cases per standard), and reported as a percentage.

### Statistical analysis

Statistical tests (Shapiro-Wilk) showed that the difference between M2 and M1 was normally distributed (W=0.983, p=0.094); therefore, paired t-tests were used to test whether there was evidence of improvement in scores from M1 to M2.

### Ethical considerations

Ethical approval was sought and obtained from both the Liverpool School of Tropical Medicine as the study sponsor (ref: 20-031) and the National Health Research Ethics Committee of Nigeria (ref: NHREC/01/01/2007). As individual consent is not normally part of the clinical audit process and no personally identifiable information was collected, consent was not sought.

## Results

Although the QI training was conducted in three countries, data from Tanzania ML and Zanzibar are presented separately as they were implemented by separate teams and showed marked differences at each stage.

Of the 128 facilities that were followed up, 91 were primary-level healthcare facilities, and 37 were either secondary- or tertiary-level facilities (Figure 3). Primary healthcare level facilities were predominant in Kenya, Nigeria, and Tanzania’s ML, but Zanzibar incorporated mainly secondary- or tertiary-level hospitals.

**Figure 3.**
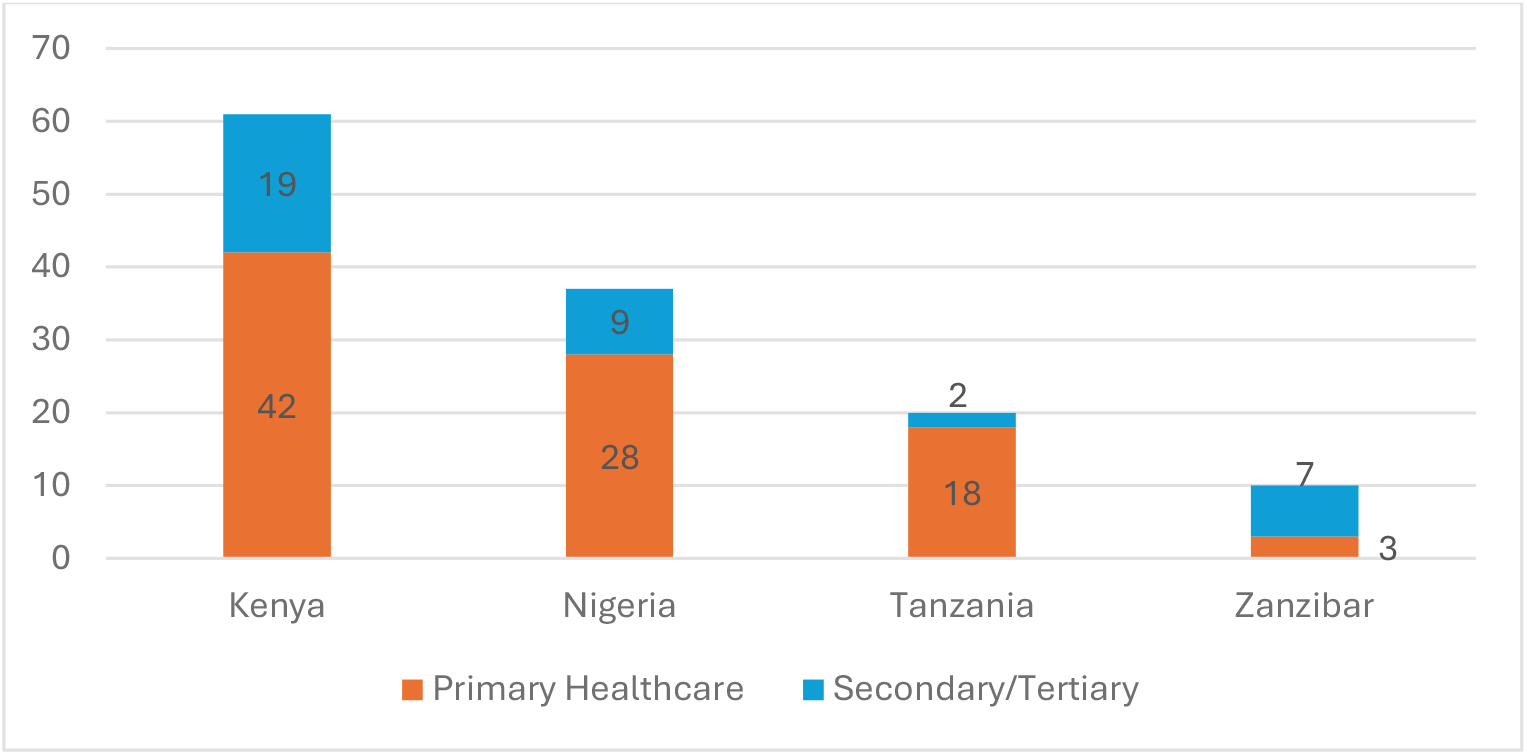
Number of health facilities by country and level

### 1. Identification

Across the three countries, all of 128 follow-up facilities appointed a QI champion, and identified an appropriate standard to audit. As facilities were encouraged to identify problems specific to their context and corresponding standards against which to measure, these varied from one site to another. Frequently selected standards were related to antenatal screening for syphilis, hepatitis B, HIV, or anaemia; healthcare workers providing respectful care; and provision of postnatal care within the first 24hours after birth.

### 2. Assessment

Overall, 96% (n=123) of the facilities conducted at least one baseline measurement (M1), calculated the percentage of instances in which the standard was met, set a target, and conducted a follow-up measurement (M2). In total, 137 audits were conducted, with target, baseline, and follow-up data collected: 61 in Kenya, 37 in Nigeria, 19 in Tanzania ML, and 20 in Zanzibar. Baseline scores ranged considerably from 0% to 93%, with a mean of 38.8% for all countries. Tanzania ML had the highest overall country M1 score at 57.3% (range: 0-93%) and Zanzibar the lowest overall score at 24.8% (range: 0-72%) (Figure 4).

**Figure 4.**
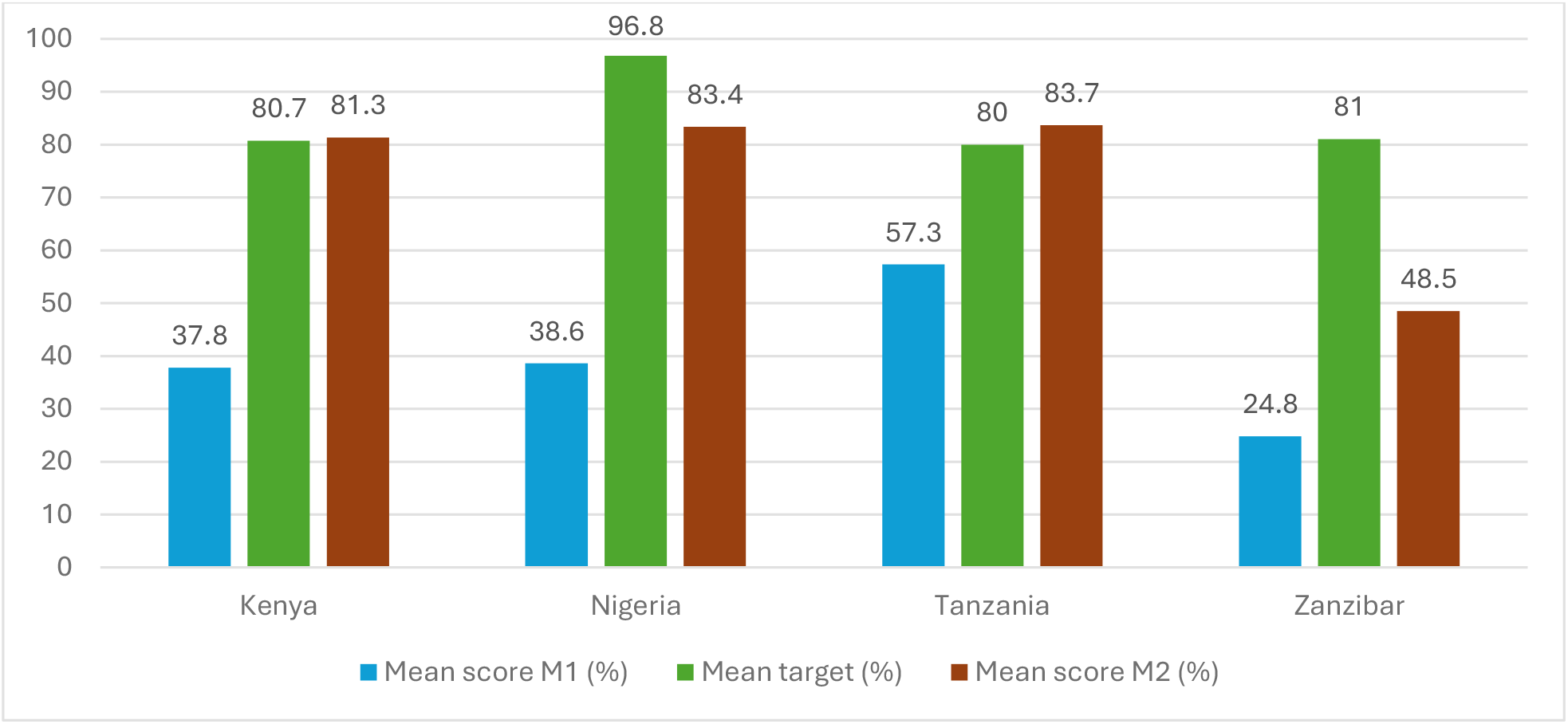
Mean percentage scores for M1, target scores, and M2 scores, by country

As part of the training, participants were encouraged to set realistic targets that were achievable while improving the quality of care provided. The mean target scores ranged from 80-96.8% (Figure 4), but there was considerable variation between individual facilities, with one facility in Zanzibar setting a target of 30% following an M1 score of 0, and several facilities in Kenya, Nigeria, and Zanzibar setting targets of 100%.

### 3. Intervention

The actions implemented as part of the StBA process were dependent on the problems identified and the specific needs of individual sites. Commonly reported actions included training or sensitising facility staff on the importance of specific aspects of care, such as screening for certain health conditions identified as problematic, including anaemia, HIV, and syphilis, and ensuring the provision of necessary items of equipment or supplies, such as haemoglobin testing kits.

### 4. Follow-up

The mean follow-up (M2) score across all countries was 77.4%, although the mean M2 scores ranged from 48.5% (range: 18-87%) in Zanzibar to 83.7% (range 28-100%) in Tanzania ML. Figure 5 shows scores at M1 compared to M2 for individual audits, with most above the blue line, indicating improvement. The overall mean difference from baseline to follow-up (M2-M1) was 38.6%, SD=23.4 (Kenya: mean=43.5%, SD=19.6; Nigeria: mean=40.7%, SD=22.7; Tanzania ML: mean=26.5%, SD=29.1; Zanzibar: mean=23.6, SD=15.2). Paired t-tests showed a statistically significant difference between M1 and M2 scores, t(136)=-19.25, p<0.001 for all countries combined, and the improvement was also highly significant for all four sites individually (p<.001).

**Figure 5.**
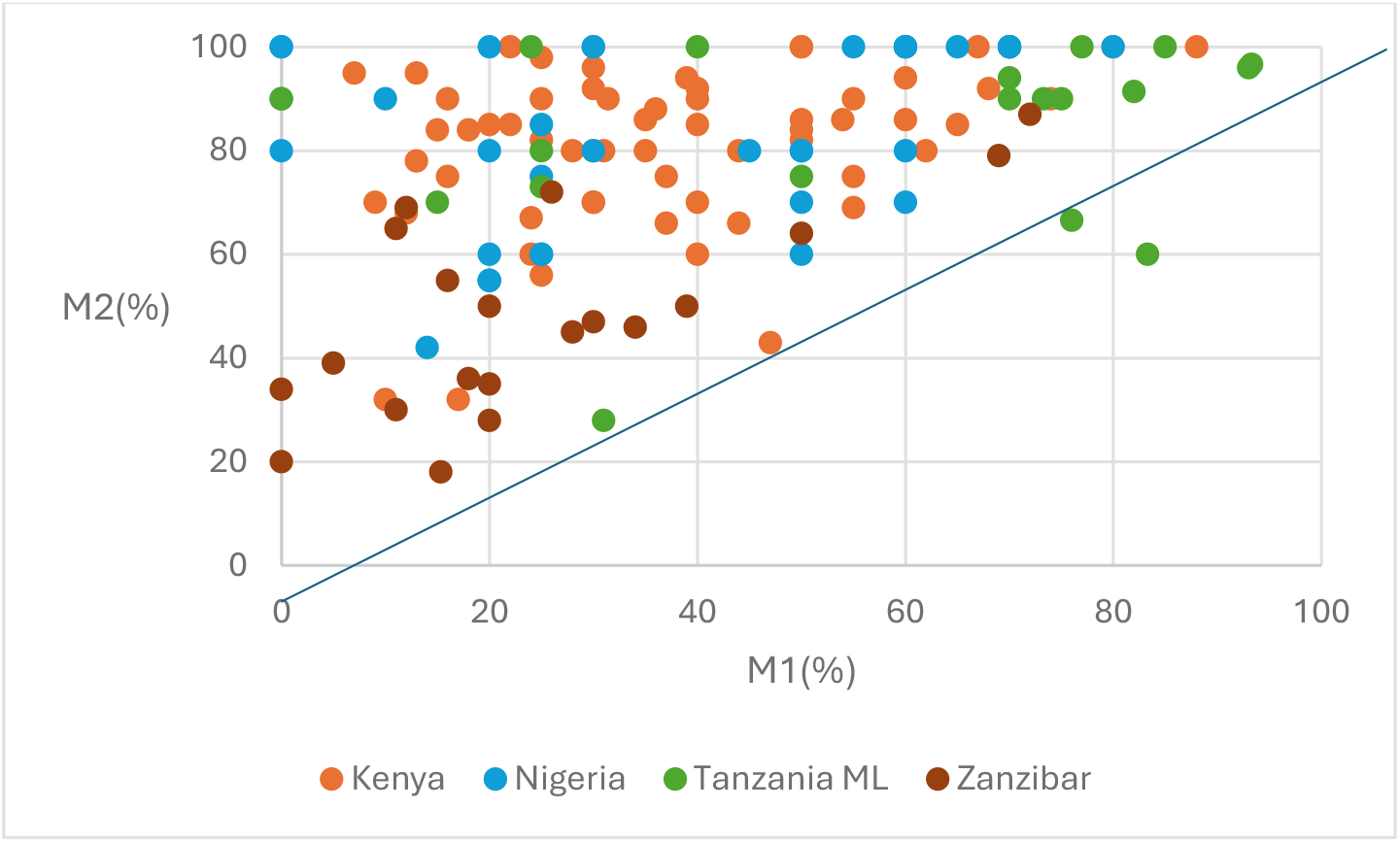
Mean scores at M1 compared to M2 by audit

Of the 137 audits conducted, in 54% (n=74) of the instances, the target score was achieved or exceeded (Figure 6). Where the standard was not achieved and information was available, most sites stated that they would continue to implement the process to further improve the standards of care provided. In the instances where a reason was given for not achieving the target, these were largely due to localised issues such as lack of staff skills/time, ongoing building renovation work, and women’s lack of awareness of targeted conditions.

**Figure 6.**
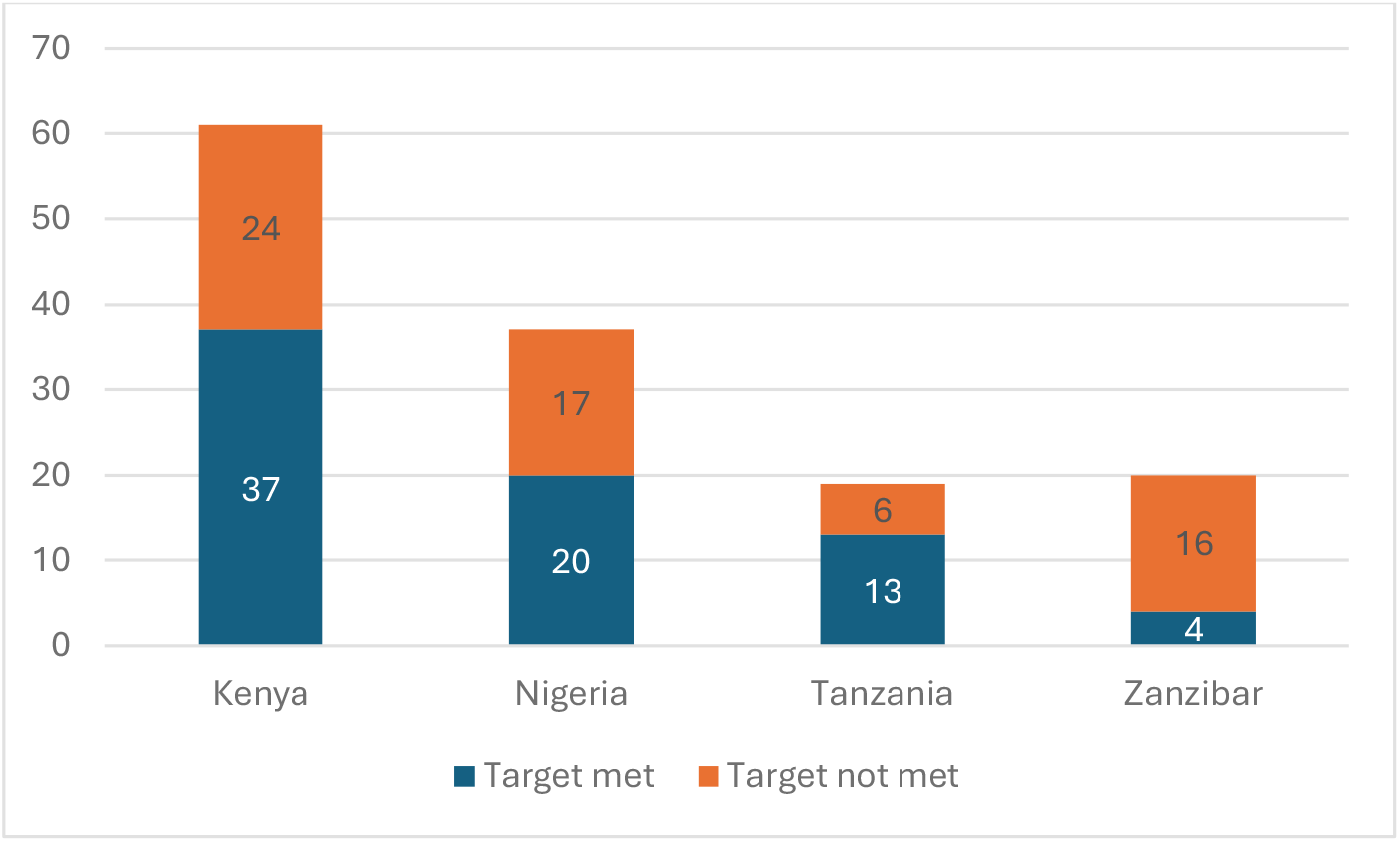
Number of audits where the target score was met

## Discussion

### Summary

Improving care quality is a key factor in reducing maternal and neonatal mortality and morbidity in LIMCs. The LSTM QI training course was deployed to nearly 900 healthcare workers, across three countries, over a four-year period. Follow-up of 128 health facilities with QI-trained staff showed good implementation of the principles of StBA and subsequent improvements in the quality of care associated with those standards. This study demonstrates that StBA can be successfully implemented in primary, secondary, and tertiary facilities in LMICs, with significant improvements in care quality. By encouraging QI teams to implement standards associated with local problems, this reduced comparability across facilities but resulted in context-sensitive interventions that were more likely to have a greater impact. This is in line with broader global health debates on the need for locally adaptive QI approaches rather than one-size-fits-all solutions (WHO, 2018). However, the lack of comparability across facilities also highlights a common trade-off in global health: balancing local ownership with the need for standardised metrics to enable benchmarking and accountability. The QI training and subsequent StBA implemented in this study can be used to address structural, process and outcome aspects the WHO Quality of Care Framework (Tuncalp et al, 2015), being particularly relevant at promoting the provision of evidence based practice, respectful care and coverage of key practices.

### National targets

While both Kenya and Tanzania’s ML exceeded the mean target scores in their facilities, Nigeria and Zanzibar did not achieve this goal. In the case of Nigeria, this may have been due to them having higher targets, as they still achieved high levels of improvement at M2, comparable to both Kenya and Tanzania ML. Zanzibar started with much lower baseline M1 scores than the other three sites but their targets were similar to those of Kenya and Tanzania ML. Their low baseline scores may have contributed to their failure to reach the targets, but they still achieved a mean level of improvement similar to that of the Tanzania ML. These findings reflect wider discussions in QI research around realistic goal-setting, equity of starting points, and the risks of performance metrics masking localised achievements (Kruk et al., 2018).

### Facility targets

Where individual facility targets were not met, discussions with facility QI teams found that in some instances, initial targets were set unrealistically high, such as setting a target of 100% following a baseline of <20%. The specific facility achieved a very good result of 90% compliance with the standard but technically still failed to reach their target. Other facilities only missed by 2-3% or needed extra time for interventions already in place, to take effect. Some standards implemented required interventions beyond the control of the staff, such as the supply of specific drugs or equipment. Where these were not obtainable due to national shortages, it resulted in failure to reach the set targets. Following the training, staff enthusiasm for the audit process led to attempts to audit multiple standards simultaneously the same time. This led to a diffused focus, and although improvements were seen, standards were not necessarily met. Standardised training and support materials were put in place, but some facilities did not always follow the prescribed audit methods.

### Systemic barriers to implementation

Several systemic barriers affected the implementation of QI training, including staff illness, lack of necessary equipment and supplies, national industrial action, and climate-related challenges such as severe flooding. These findings reinforce the argument that QI cannot just be isolated at the facility level but must be situated within the broader health system and its enabling environment (Frenk, 2010).

The implementation of an intervention targeting a standard with an already high baseline of 93% severely limited the potential for measurable improvement. This raises concerns about the selection process for target problems and standards and suggests the potential for management or administrative influence aimed at presenting the facility in a more favourably.

Resource constraints, such as lack of staffing, and supplies of necessary drugs and equipment, highlight the systemic nature of QI. Successful and sustainable QI initiatives require an enabling environment, with strong governance and leadership, adequate and predictable financing, efficient supply chains, and effective health information systems. Furthermore, health systems need to be resilient to external shocks, such as extreme weather events which can severely disrupt service delivery. Ensuring continuity in the supply of critical commodities requires strategic planning and the development of contingency plans.

These challenges highlight the importance of capacity building, supportive supervision, and leadership in embedding QI approaches (Mosadeghrad et al., 2023). They are linked to the global debate on how to build a culture of quality in health systems, balancing technical methods with behavioural and organisational changes.

This study found that standards-based audits can be successfully implemented in facilities of all levels with positive results. It can be used as a tool to improve care quality and associated maternal and newborn outcomes. It is a feasible, low-tech approach which can be trained using blended learning methods but can be constrained by the lack of system enablers, including adequate staffing, good governance, and effective supply chains. To promote sustainability and scalability, QI methods need to be moved from a facility-level activity to a more health systems approach (Waiswa, 2020).

### Study limitations

Although this study produced significant improvements in the targeted quality of care, the sites included in Kenya and Nigeria were selected by local and national authorities based on patient turnover and were located in states/counties already receiving quality improvement (QI) interventions. These interventions included staff training in antenatal and postnatal care, mentorship, and supportive supervision. The study sites in Tanzania were randomly selected but were still stratified by geographical location in a QI implementation region. Other limitations of the study were that health facilities were only followed up for a relatively short period of time (3-6 months) and that there was no assessment of the impact of the interventions on maternal and neonatal outcomes or cost-effectiveness. There was also the potential risk of the Hawthorne Effect (McCambridge et al, 2014), the awareness by participants that they are being studied, might have affected their practice, over and above the effect of the intervention.

### Recommendations for overcoming implementation challenges

Future research into the cost-effectiveness of QI training could compare the cost of implementing the programme versus improvements in health outcomes, supporting its advocacy to regional and national governments. The use of randomised or quasi-experimental and mixed-methods study designs would improve the generalisability of the evidence and provide a more rounded evaluation of the impact of StBA, including from the service user perspective. Additionally, longer term follow-up would allow the assessment of the health outcomes of women and newborns, before and after implementation, and mapping of interventions against the WHO recommendations for quality health services (WHO, 2025b)

## Conclusion

Implementing StBA following QI training is associated with substantial improvements in audited care processes across varied settings. This study showed an increase of over a third in the scores of audited standards, with more than half of the facilities achieving their targets. The contextualisation of the selection of standards allows for interventions to be more relevant to local settings and maximises the potential for impact; however, incorporating the process into health systems could promote its scale-up and sustainability. Future research should test the durability and cost-effectiveness of the intervention, and subsequent patient outcomes.

## Data Availability

All data produced in the present study are available upon reasonable request to the authors

## Acknowledgements

The authors wish to acknowledge the support of Adacha Boslam Bello (Nigeria), Rael Mutai (Kenya & Tanzania), Lucy Nyaga (Kenya), Lalashe Kiretun, Shuwena Hamad, Mansab Ramadhan Mansab, Maria Angelica Rweyemamu, Zainab Suleiman (Tanzania/Zanzibar), Sarah White (UK), and programme support teams in all three countries. The authors also wish to thank the Ministries of Health, Reproductive Health Co-ordinators, and healthcare professionals in each of the three countries for their support of the training implementation and data collection; and to Global Fund and Takeda Pharmaceutical Co. for funding the study.

## Ethical approval

Ethical approval for this study was obtained from Research Ethics Committees at, Liverpool School of Tropical Medicine (ID:21–052), Nigeria (Ministry of Health Oyo State: AD13/479/44511), Kenya (NACOSTI/P/21/13853), Tanzania (University of Dodoma: MA.84/261/02/‘A‘/25 and Zanzibar Health Research Institute: ZAHREC/04/PR/JUNE/2022/19).

